# Testing effectiveness and implementation of a standardized approach to sexual dysfunction screening among adolescent and young adult-aged survivors of childhood cancer: A type I hybrid, mixed methods trial protocol

**DOI:** 10.1101/2024.06.05.24308510

**Authors:** Jenna Demedis, Julia Reedy, Kristen Miller, Junxiao Hu, James L. Klosky, Brooke Dorsey Holliman, Pamela N. Peterson, Eric J. Chow, Christina Studts

## Abstract

**Background:** Approximately 20-50% of adolescent and young adult-aged childhood cancer survivors (AYA-CCS) experience sexual dysfunction (SD), although this healthcare need is widely underrecognized. Previous research from both AYA-CCS patients and their providers report that SD needs are unaddressed despite patient desires for SD discussions to be incorporated as part of their care. Patients and providers agree that standardized use of a patient-reported outcome measure may facilitate SD discussions; an SD screening approach was developed with patient and provider input. This study will measure the effectiveness of a standardized SD screening intervention and assess implementation outcomes and multilevel barriers and facilitators to guide future research.

**Methods:** This multi-site, mixed methods, type 1 effectiveness-implementation hybrid trial will be evaluated using a pre-post design (NCT05524610). The trial will enroll 86 AYA-CCS (ages 15-39) from two cancer centers in the United States. The SD intervention consists of core fundamental functions with a “menu” of intervention options to allow for flexibility in delivery and tailoring in variable contexts. Effectiveness of the intervention on facilitating SD communication will be measured through patient surveys and clinical data; multivariable logistic regression will be used for the binary outcome of self-reported SD screening, controlling for patient-level predictors. Implementation outcomes will be assessed using mixed methods (electronic health record abstraction, patient and provider surveys, and provider interviews. Quantitative and qualitative findings will be merged using a joint display to understand factors affecting intervention success.

**Implications:** Identification and treatment of SD in AYA-CCS is an important and challenging quality of life concern. The type 1 hybrid design will facilitate rapid translation from research to practice by testing the effects of the intervention while simultaneously identifying multilevel barriers and facilitators to real-world implementation. This approach will inform future testing and dissemination of the SD screening intervention.

## Introduction

Sexual dysfunction (SD) is estimated to occur in 20-50% of adolescent and young adult-aged childhood cancer survivors (AYA-CCS) but is widely underrecognized [1–8]. SD encompasses lack of desire for sex, arousal difficulties (erection, lubrication), inability to achieve climax/ejaculation, anxiety about sexual performance, climaxing/ejaculating too rapidly, physical pain during intercourse, and lack of pleasure [9]. In AYA-CCS, SD may occur as a result of physiologic and psychosexual reasons, and while certain treatment factors (e.g., cranial or pelvic irradiation, central nervous system tumors, chemotherapy) exist, demographic, developmental, psychologic and social factors also contribute [1–8, 10–13]. SD is associated with poorer quality of life, including physical, social, and mental health; therefore, providing assessment, education, and counseling for AYA-CCS experiencing SD is critical [1–4, 9–12].

While data are limited in cancer patients of all ages generally, two studies have suggested that addressing SD may improve psychological distress [14, 15]. Because the multidisciplinary pathophysiology of SD in this population makes estimating individual SD risk difficult, all AYA-CCS warrant SD-focused education and evaluation.

Despite the prevalence and impact of SD among AYA-CCS, patients largely report that their needs in this area are unaddressed by their primary care providers and oncologists [7, 16–19]. This is consistent with findings from providers, with 50% of pediatric oncologists reporting rarely or never discussing sexual function with their AYA-CCS [20]. Patient- and provider-level barriers are well described, including discomfort, lack of provider knowledge, and lack of patient understanding of how to broach the topic [19, 21–23]. Multiple qualitative studies have demonstrated that patients would like their providers to initiate conversations to address SD directly, privately and regularly [16, 19, 23]. Further, national guidelines recommend regular discussions of sexuality and cancer throughout cancer care and follow-up [24–26].

Given the aforementioned barriers, which may be particularly salient in younger populations, our prior research evaluated the utility of using a patient-reported outcome measure for SD screening in AYA-CCS followed in pediatric settings (age 15-24 years), eliciting patient and provider perspectives in qualitative interviews [22, 27]. Both groups were in favor of using a patient-reported outcome measure to address barriers and facilitate SD conversations. While no SD screening tools have been specifically validated in the AYA-CCS population, our prior research qualitatively evaluated the National Institute of Health-developed Patient-Reported Outcomes Measurement Information System (PROMIS®) Sexual Function and Satisfaction (SexFS) Brief v2.0 tool in the AYA-CCS population [27, 28]. The SexFS Brief was found to be acceptable and useful, with demonstrated response process and content validity [27]. In addition to evaluating the utility of the SexFS Brief, patients and providers offered perspectives for how the SexFS Brief could be implemented into clinical use, including recommendations for delivery, results storage and review, and needed resources [22, 27].

These recommendations were included in an SD screening intervention prototype that is being iteratively adapted in an ongoing phase of research. Once iterative adaptations are completed, the study described here will simultaneously test effectiveness and implementation outcomes of this screening intervention.

Clinical use of the SexFS Brief has the potential to improve screening and detection of SD among AYA-CCS, which is the first step toward providing SD-related healthcare and ultimately improving related quality of life concerns. However, because SD screening utilizing the SexFS Brief is not yet part of routine care with the AYA-CCS population, and because patient-reported outcomes are not commonly used in this population, additional research is required to evaluate the effectiveness of the screening intervention in facilitating SD communication, as well as its potential for implementation in clinical settings.

## Study Purpose

The purpose of this pilot type 1 hybrid effectiveness-implementation trial is to 1) demonstrate preliminary effectiveness of the standardized SD screening intervention in improving SD communication between AYA-CCS (current ages 15-24 years) and their providers, and to 2) assess implementation outcomes and multilevel barriers and facilitators to guide future research. Use of a hybrid trial design will facilitate rapid translation from research to practice by addressing implementation, service, and client outcomes [29]. This study will use mixed methods to quantitatively measure effectiveness and implementation outcomes in a large sample while simultaneously contextualizing these outcomes by integrating them with complementary in-depth qualitative data analysis. Quantitative data at the patient and provider levels will be obtained through surveys and from electronic health record (EHR) data.

Quantitative data will be supplemented with qualitative data collection via concurrent open-ended survey questions as well as nested sequential explanatory qualitative interviews with providers. The following quantitative, qualitative, and mixed methods research questions will guide the study:

### Quantitative

#### Effectiveness

What effects does the SD screening intervention have on patient-reported SD communication and patient satisfaction with SD healthcare? What impact does the intervention have on clinical care (e.g., documentation of SD, referral patterns)?

#### Implementation

How do patients and providers rate the SD screening intervention’s preliminary implementation outcomes (acceptability, appropriateness, feasibility and fidelity)? How often is the intervention utilized by patients and providers?

### Qualitative

#### Effectiveness

What are providers’ impressions of the SD screening interventions’ effectiveness? What are providers’ perceptions of barriers and facilitators affecting the interventions’ effectiveness?

#### Implementation

What are the barriers and facilitators to adopting and implementing the SD screening intervention? How does the intervention fit in a pediatric oncology clinic context? Are there other contextual factors that affect its use and utility? Are changes to the intervention needed?

### Mixed Methods

#### Effectiveness

What barriers and facilitators exist that may explain or expand upon effectiveness results, including variability in clinical effects? What changes to the SD screening intervention could improve its effectiveness?

#### Implementation

What contextual factors are contributing to the SD screening intervention’s implementation outcomes? What changes to the intervention could improve its reach, acceptability, feasibility, and other implementation/process outcomes?

## Methods and analysis

### Study Frameworks

The overall structure and outcomes of this study are guided by the implementation science framework, RE-AIM (Reach, Effectiveness, Adoption, Implementation and Maintenance) [30]. RE-AIM is used to systematically assess multiple outcomes of intervention implementation, including effectiveness, with the overarching goal of increasing the population impact of interventions through widespread adoption, implementation, and maintenance reaching all patients who could benefit. In this study, RE-AIM will inform our assessment of screening intervention effectiveness, reach, adoption and implementation across settings and patient subgroups, as well as its potential for scaling up and spreading to additional settings.

While RE-AIM will guide planning and evaluation of effectiveness and implementation, the updated Consolidated Framework for Implementation Research (CFIR 2.0) will inform evaluation of contextual factors, including multilevel barriers and facilitators, that may influence RE-AIM domains, informing future intervention adaptation and implementation [30–36]. Use of RE-AIM and CFIR 2.0 is further described in the Data Collection section below.

### Study Design

A multi-site type 1 effectiveness-implementation hybrid trial design will be used to simultaneously establish preliminary effectiveness and implementation outcomes (Table 1; Figure 1; S1 Protocol). As a type 1 hybrid trial, the primary objective of this study is to determine the effectiveness of the SD screening intervention at improving the occurrence SD-focused patient-provider communication. Additional effectiveness and implementation outcomes will be secondary/exploratory and are detailed below.

**Figure 1.**
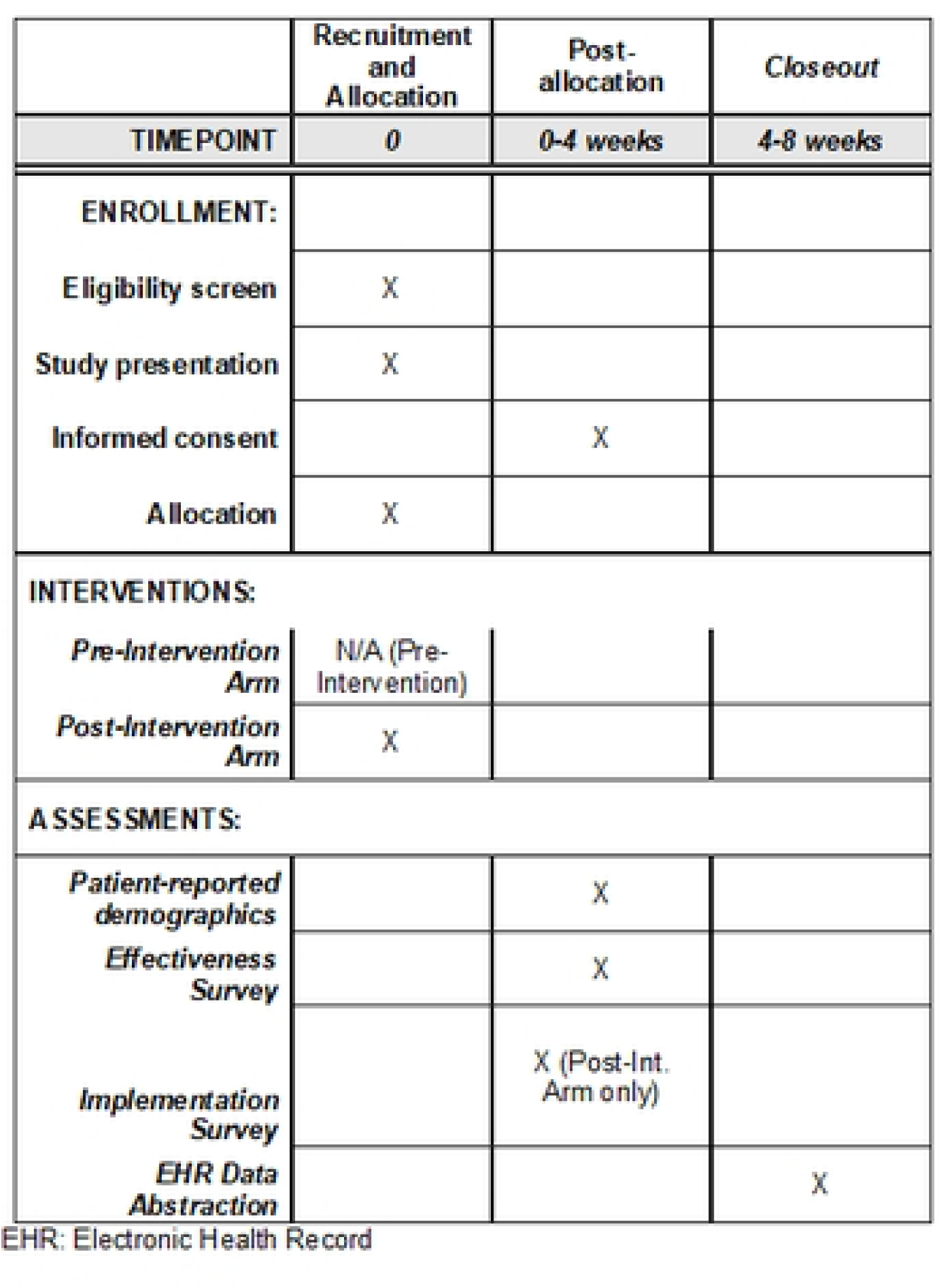
Schedule of patient participant enrollment, screening intervention, and assessment.

**Table 1.**
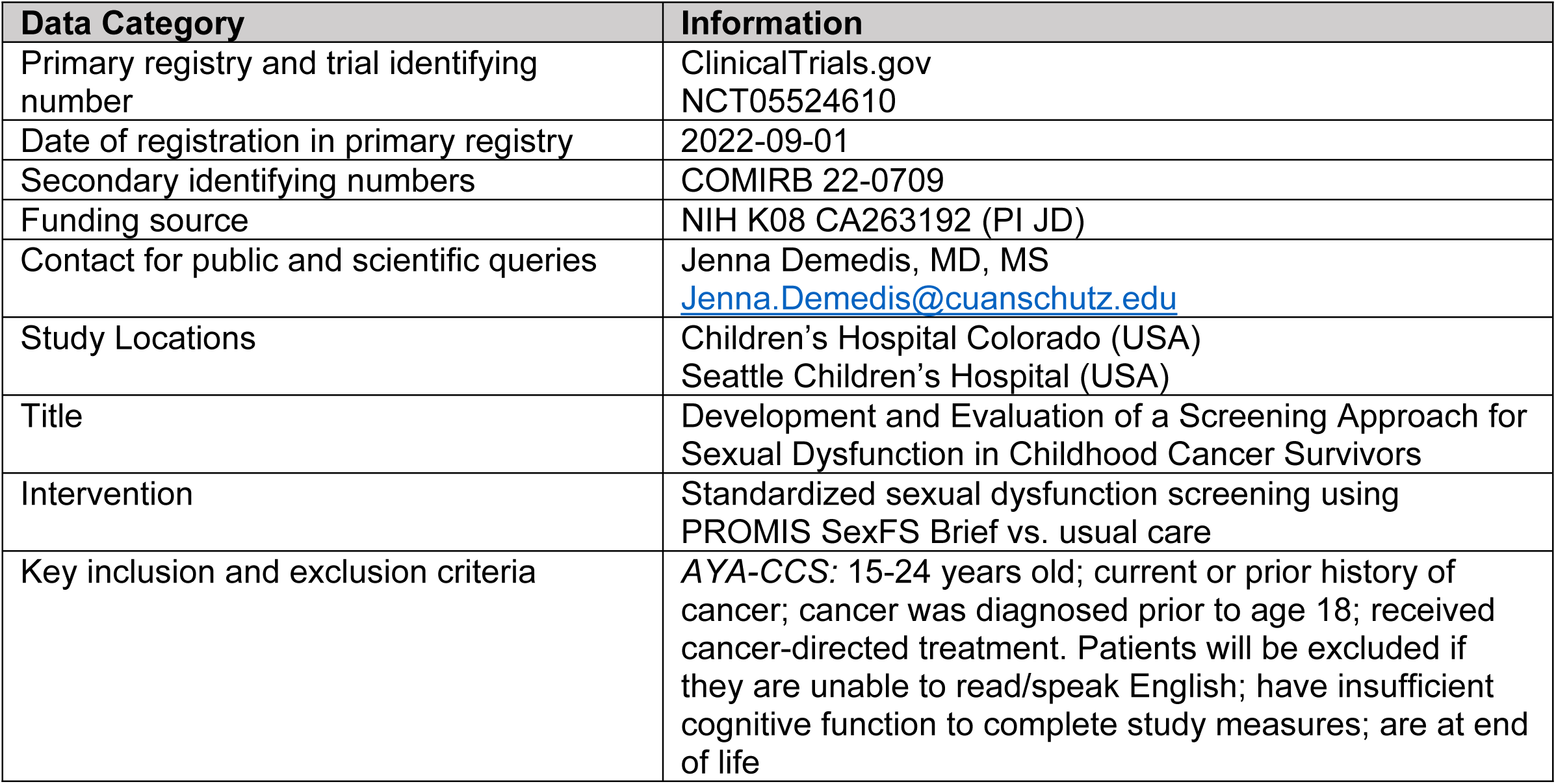
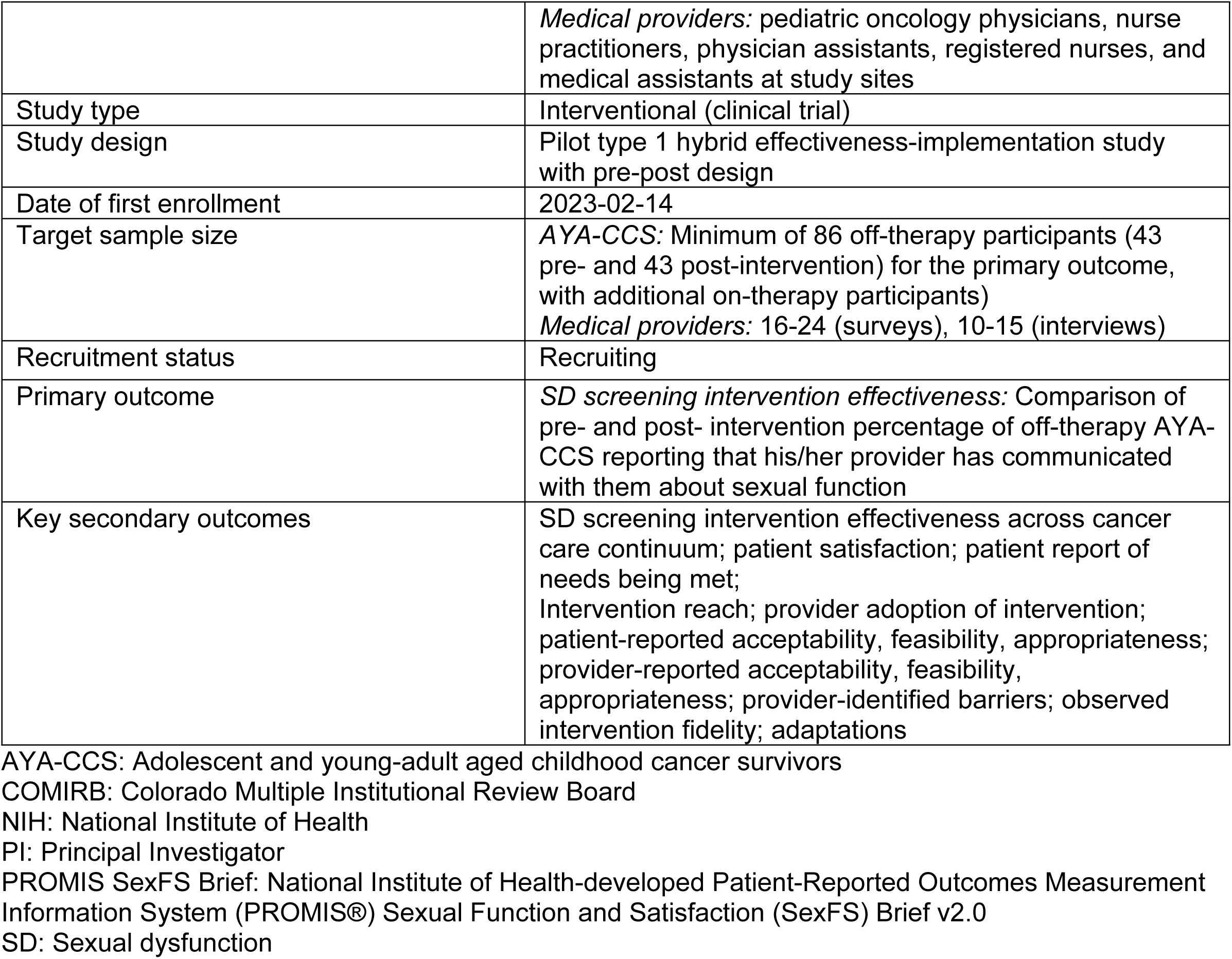
Protocol Overview.

|  |  |
| --- | --- |
|  | *Medical providers:* pediatric oncology physicians, nurse practitioners, physician assistants, registered nurses, and medical assistants at study sites |
| Study type | Interventional (clinical trial) |
| Study design | Pilot type 1 hybrid effectiveness-implementation study with pre-post design |
| Date of first enrollment | 2023-02-14 |
| Target sample size | AYA-CCS: Minimum of 86 off-therapy participants (43 pre- and 43 post-intervention) for the primary outcome, with additional on-therapy participants)<br>*Medical providers:* 16-24 (surveys), 10-15 (interviews) |
| Recruitment status | Recruiting |
| Primary outcome | *SD screening intervention effectiveness:* Comparison of pre- and post- intervention percentage of off-therapy AYA-CCS reporting that his/her provider has communicated with them about sexual function |
| Key secondary outcomes | SD screening intervention effectiveness across cancer care continuum; patient satisfaction; patient report of needs being met;<br>Intervention reach; provider adoption of intervention; patient-reported acceptability, feasibility, appropriateness; provider-reported acceptability, feasibility, appropriateness; provider-identified barriers; observed intervention fidelity; adaptations |

AYA-CCS: Adolescent and young-adult aged childhood cancer survivors
COMIRB: Colorado Multiple Institutional Review Board
NIH: National Institute of Health
PI: Principal Investigator
PROMIS SexFS Brief: National Institute of Health-developed Patient-Reported Outcomes Measurement Information System (PROMIS®) Sexual Function and Satisfaction (SexFS) Brief v2.0
SD: Sexual dysfunction

The standardized SD screening intervention will be tested in a pre-post study design in which the SD screening intervention will be implemented with all patients as standard of care; this is justified by national guidelines recommending universal sexual health conversations for all age-appropriate oncology patients. Prior to initiating the SD screening intervention, patient surveys will be used to collect baseline data on SD communication and clinical care.

Subsequently, the SD screening intervention will begin. After intervention initiation, quantitative (survey, EHR) and qualitative (interviews, open-ended questions) data will be collected to evaluate effectiveness and implementation outcomes.

#### Intervention Design and Implementation Package

The standardized SD screening intervention is a multicomponent intervention composed of required components with flexible delivery, tailored to each study site based on a “menu” of options (Table 2). This approach follows the implementation science concept of functions and forms of an intervention: functions are the fundamental purpose or desired effect of the intervention, while forms are the intervention details, modalities, and activities that may vary based on contextual factors [37, 38]. The core functions of the SD screening intervention, detailed in Table 2, include (1) routine/standardized SD screening using the SexFS Brief, (2) consistent follow-up and management of screening results, and (3) patient and provider education. These functions address the key barriers driving inadequate SD-focused healthcare: AYA patients’ uncertainty about SD risks and ability to address these with the medical team, and clinicians’ lack of awareness of SD risks and how to address/manage these. Examples of the “forms” that each function may take, such as details of the target population or modality of delivery, are detailed in Table 2. At study initiation, the research team will work with each clinical site to adapt the SD screening intervention to their context, selecting from the “menu” of forms. The research team will provide relevant background information from prior phases of research (e.g., patient- and provider-recommendations and preferences) to inform tailoring to each site’s unique context, including existing resources, workflows and barriers.

**Table 2.** Core functions and forms of the sexual function screening intervention.

| PATIENT LEVEL |  |  |
| --- | --- | --- |
| MOTIVATING NEED/PROBLEM | CORE FUNCTIONS | FORMS - DECISIONS AND EXAMPLES |
| AYA oncology patients are uncertain of sexual dysfunction (SD) risks or ability to address them with their medical team | Patient knowledge of SD risks is increased through education | I. Patient Education - Examples:<br>· Primary team educates patients during treatment discussion with fertility education<br>· Standard patient education sessions (e.g. Oncology Class) includes section on sexual health/function<br>· Reminders and re-introduction with each screening event<br>· Non-in-person modalities: video, pamphlets or other reading materials, website |
|  | Patients participate in regular written screening for SD concerns using a patient-reported outcome measure (e.g. PROMIS SexFS Brief) | II. Target population - Decisions and Examples:<br>· Age of inclusion (e.g. 12 years of age and older, 15 years of age and older)<br>· Treatment status: patients actively receiving therapy, survivors only<br>· Patient choice: Opt out vs. opt-in |
|  |  | III. Screening delivery - Decisions and Examples:<br>· Technology-based (e.g. through medical record, separate website) delivery (e.g. tablet/phone in clinic, text, email, mobile application)<br>· Paper-based delivery (e.g. in clinic, mailed)<br>· Non-English readers (translate tool, deliver verbally)<br>· Non-readers (omit, deliver verbally) |
|  | Patients are enabled to address SD concerns through increased comfort and privacy | IV. Patient Privacy and Comfort - Decisions and Examples:<br>· Location of screening (e.g. at home, in clinic waiting room, in clinic private room)<br>· Timing of screening (e.g. before visit, during rooming of patient, while waiting for provider or other services, at end of visit)<br>· Minimizing parent/guardian presence during screening and conversations (asking parents to step out of room, providing separate space for screening or education, bringing patients back separately for screening)<br>· Consideration of patient choice in who addresses SF concerns (e.g., specific member of team, provider of same sex, etc.) |
| PROVIDER LEVEL |  |  |
| MOTIVATING NEED/PROBLEM | CORE FUNCTIONS (Standardized) | FORMS (Tailored) |
| Clinicians are | Increase | V. Provider Education - Decisions and Examples: |
| unaware of AYA oncology patients' SD needs and how to address them | provider awareness, knowledge and comfort of SD needs | · *Modality of education (e.g. written education such as handouts/powerpoints or interactive education such as computer modules/lectures/meetings)*<br>· *Target provider population (e.g. physicians, advanced practice providers, nurses, medical assistants)*<br>· *Amount of education provided to each provider (e.g. universal materials, or graduated depending on role in clinic or role within sexual health care)*<br>· *Timing/frequency of provider education (e.g. single time point, regular interval re-education/reminders, performance-based education)* |
|  | Consistently and periodically screen for SD needs using the PROMIS SexFS Brief | VI. Screening Timing - Examples<br>· *Initiation of screening (e.g. upon diagnosis, 1 month after diagnosis, at completion of therapy)*<br>· *Screening frequency: single time point, systematic interval (e.g., every 3 months or as frequently as patient comes) or patient tailored (e.g., frequency depends on diagnosis, patients' sexual history, etc.)*<br>VII. Consistency and Sustainability - Decisions and Examples<br>· *Engaged personnel: established clinical champion(s), dedicated team, obtaining provider input/feedback on implementation*<br>· *Clear process for triggering screening (e.g. systematizing screening (automatic in medical record) or dedicated care manager/role)*<br>· *Provider reminders (e.g. in medical record manually or automatized via EHR functionality), on chemotherapy roadmaps)*<br>· *Provider feedback (e.g. regular updates on screening rates, notification of missed screening)* |
|  | Consistently address SD needs once identified | VIII. Management of screening results - Decisions and Examples<br>· *Results storage modality: electronic (e.g. medical record, other database), paper (patient charts), or omit longitudinal storage)*<br>· *Results/management storage and privacy: secure notes for minors vs. avoidance of documentation vs. normal documentation, limited vs. unrestricted access to results by medical providers*<br>· *Follow-up of screening results with patient: conversation with patient (in person, on phone, electronic), provide materials to patient (pamphlet/materials/website), automatic referral to specialists*<br>· *Responsible providers: existing oncology team, internal dedicated care manager or team, external specialist*<br>· *Management resources: evidence-based clinical decision-making tools, referral guides, established specialist partnerships* |

PROMIS SexFS Brief: National Institute of Health-developed Patient-Reported Outcomes Measurement Information System (PROMIS®) Sexual Function and Satisfaction (SexFS) Brief v2.0 SD: Sexual dysfunction

Similarly, strategies for implementing the SD screening intervention, such as the plan for provider education and support, will be tailored by site (Table 3). The intervention and implementation menus were developed based on prior patient- and provider-engaged stages of research. Further, the implementation menu was informed using established implementation science strategies to target known barriers [19, 22, 39]. For example, to address lack of provider knowledge, implementation should include provider education and resources, which can take several forms including education sessions, written education, clinical decision-making tools, or development of a dedicated expert team. Each strategy is intended to target study outcomes including effectiveness, adoption, and implementation (acceptability, appropriateness).

**Table 3.** Implementation strategies.

| Component | Sub-component | Options | Outcome Target |
| --- | --- | --- | --- |
| Intervention design | Planning | -Selection of key partners (providers, patients, clinical leadership, RNs, MAs) in selecting flexible intervention components and implementation plan | Effectiveness<br>Reach<br>Adoption<br>Implementation (Acceptability, feasibility) |
|  | Iteration/Scale-up/Stage | -Optional stage |  |
| Provider education | Modality | -Emails<br>-Written education document<br>-Presentations/meetings<br>-Online modules | Effectiveness<br>Adoption<br>Implementation (acceptability, appropriateness) |
|  | Content | -In depth vs. Overview |  |
|  | Who | -Only dedicated team members<br>-All providers |  |
| Provider Resources | Written | -Evidence-based clinical decision-making tools<br>-Referral guides/work-up flow diagram | Effectiveness<br>Reach<br>Adoption<br>Implementation (acceptability, appropriateness) |
|  | Personnel | -Development of clinical team<br>-Established specialist partnerships |  |
| Reminders | Written | -EHR-based questionnaire automatic triggers<br>-Dedicated team monitors at patient-level<br>-EHR-Based (Best Practice Alerts, notes in chart)<br>-Paper based/existing clinic documents (roadmaps, care pathways) | Reach<br>Adoption<br>Implementation (fidelity) (Sustainability) |
|  | Verbal | -Clinical champions |  |
|  | Auditing/feedback | -Regular updates on screening rates<br>-Notification of missed screening |  |

EHR: electronic health record
MA: medical assistant
RN: Registered nurse

### Study Setting and Population

This trial will occur in four clinical settings at two academic stand-alone children’s hospitals: Children’s Hospital Colorado and Seattle Children’s Hospital.

Both patients and medical providers will be recruited for participation. Patients will be eligible for the study if they are 1) 15-24 years old at time of enrollment; 2) Have a current or prior history of cancer (International Classification of Diseases for Oncology (ICD-O) with behavior code ≥2); 3) Cancer was diagnosed prior to age 18; and 4) Have received cancer-directed treatment (chemotherapy, immunotherapy, radiotherapy, or partial/total resection). Due to the nature of study procedures, patients will be excluded if they are 1) Unable to read/speak English; 2) Have insufficient cognitive function to complete study measures; 3) At end of life.

Further, patients who participated in intervention development will be excluded and participants who do not receive the intervention will not answer implementation-focused survey questions. Of note, while AYA-CCS both receiving and having completed therapy will be included for secondary and exploratory outcomes, the primary outcome will evaluate effectiveness of the intervention among participants who have completed therapy (“off-therapy”). The population for the primary outcome was designed to mirror the inclusion criteria of our preceding qualitative study; the population was expanded to include patients currently on-therapy for secondary analyses based on subsequent recommendations from providers [22, 40].

Importantly, while the study will only include patients who can read and speak English because the SexFS has only been validated in English, the clinical intervention will still be available to non-English speakers, through verbal screening delivery via an interpreter.

All providers caring for patients at the study sites, including physicians, nurse practitioners, physician assistants, registered nurses, and medical assistants, will be eligible for participation.

#### Sample Size

As a type 1 hybrid effectiveness-implementation trial, this study is powered to detect preliminary effectiveness of the SD screening intervention. The primary outcome is the proportion of off-therapy AYA-CCS participants reporting SD conversations with their providers (Table 4); on-therapy patients are included in this study as an exploratory aim but are not included in minimal sample size based on power calculation. To detect a clinically relevant difference of 25% (5% pre-intervention vs. 30% post-intervention) in SD conversations with a multivariable logistic regression assuming 20% of variability is explained by other predictors, 80% power, and alpha =.05, 86 participants will be required (43 pre- and 43 post-implementation of screening approach) [7, 19, 23].

**Table 4.** RE-AIM outcome measurement.

| RE-AIM Domain | Outcome/Measure(s) | Method |
| --- | --- | --- |
| Reach | Proportion of eligible patients who completed SD screening | EHR abstraction |
|  | Reason for missed screening (when relevant) | Surveys (providers) |
|  | Representativeness: comparison of sociodemographic characteristics of patients receiving/not receiving intervention | EHR abstraction |
| Effectiveness<br>(primary outcome) | Primary: off-therapy AYA-CCS-report of SD communication | Surveys (patients) |
|  | Secondary:<br>All participant-report and on-therapy participant report of SD communication<br>Patient satisfaction<br>Patient-report of needs being met | Surveys (patients) |
|  | Exploratory:<br>Incidence of documented SD<br>Incidence of referral to SD-related specialty care | EHR abstraction |
| Adoption | Provider-reported incidence of viewing SD screening results | Surveys (providers) |
|  | Provider-reported incidence of discussing SD results | Surveys (providers) |
|  | Solicitation of description of adoption, explanations for quantitative findings | Interviews (providers) |
|  | <u>Exploratory:</u><br>Objective survey viewing data (e.g., EHR clicks) | EHR abstraction |
| Implementation | <u>Acceptability:</u> Acceptability of Implementation Measure (AIM) | Surveys (patients, providers)<br>Interviews (providers) |
|  | <u>Appropriateness:</u> Implementation Appropriateness Measure (IAM) | Surveys (patients, providers)<br>Interviews (providers) |
|  | <u>Feasibility:</u> Feasibility of Intervention Measure (FIM) | Surveys (patients, providers)<br>Field notes (research team)<br>Interviews (providers) |
|  | <u>Fidelity:</u> extent to which each core function, and site-adapted form, was followed | Fidelity checklist (patient-report, research team observation sampling) |
|  | <u>Adaptation:</u> intervention and implementation adaptation details tracked following FRAME and FRAME-IS | Observation, research/clinical team communication |
|  | <u>Contextual factors:</u> barriers, facilitators, recommendations | Open-ended survey questions (patients, providers)<br>Field notes (research team)<br>Interviews (providers) |
| Maintenance | Continuation beyond study |  |

AIM: Acceptability of Implementation Measure
AYA-CCS: Adolescent and young-adult aged childhood cancer survivors
EHR: electronic health record
FIM: Feasibility of Intervention Measure (FIM)
FRAME: Framework for Reporting Adaptations and Modifications-Expanded
FRAME-IS: Framework for Reporting Adaptations and Modifications to Evidence-based Implementation Strategies
IAM: Implementation Appropriateness Measure
RE-AIM: Reach, Effectiveness, Adoption, Implementation and Maintenance
SD: Sexual dysfunction

This study will secondarily explore preliminary implementation outcomes through patient implementation surveys administered in conjunction with assessment of effectiveness (n=43 off-therapy AYA-CCS, with additional on-therapy participants). Provider implementation surveys will be administered to consenting providers (anticipating an 80% response rate, approx. n=16-24), and semi-structured interviews with be completed with 10-15 providers directly involved in the SD screening intervention. Additional brief interviews may be performed to capture experiences of providers affected by, but not directly involved in, the intervention delivery (n=5-10). Because interviews will be focused on experiences with the screening approach and barriers/facilitators, we anticipate reaching thematic saturation with these sample sizes [41, 42].

#### Recruitment

Recruitment will occur on a rolling basis during existing clinic visits or via phone within 4 weeks of the existing appointment. Sampling will be purposive to achieve variation across age categories (age 15-19 and 20-24), patient treatment status (on- vs. off-therapy), and gender. In the pre-intervention phase, patient participants will be given the opportunity to consent and complete the survey at the same visit. However, post-intervention, consent and survey completion will be delayed up to 4 weeks after the patient is “due” for the SD screening intervention, to allow for buffer time for both screening and follow-up by the clinical team.

Participation will be compensated with a gift card totaling $10 pre-intervention (effectiveness survey only) and $20 post-intervention (effectiveness and implementation surveys).

Provider participants will be recruited for surveys via a variety of methods, including email or in person requests. Enrollment of providers will occur at the end of the post-implementation study phase, with all eligible medical stakeholders approached for participation in surveys. However, for sites that choose to have a limited number of providers responsible for survey delivery and follow-up/management, only those responsible providers will receive complete surveys, with other providers completing abbreviated surveys. Medical stakeholders will receive $10 gift card compensation for survey completion if the local site allows provider incentives. A subset of providers will be recruited to complete qualitative, semistructured interviews; recruitment will be purposeful to include provider participants with a range of survey responses as well as proportional representation across roles and clinics. Interview participants will receive a $40 gift card as compensation if the local site allows provider incentives.

### Data Collection

RE-AIM outcome measurement for this study is detailed in Table 4 and below. Basic demographic data, including gender identity, will be collected for all patient participants, and details about providers’ roles and experience will be collected for all provider participants.

#### Effectiveness

Effectiveness will be measured quantitatively via patient surveys (S2 Appendix). The primary outcome is a patient-reported indicator that his/her provider has communicated with them about SD. Secondary outcomes include patient satisfaction with SD communication [43] and patient-report that an SD need was met. This study will also assess the feasibility of collecting EHR data in all eligible patients via retrospective chart review, specifically collecting exploratory outcomes: 1) incidence of documented/detected SD; and 2) referral patterns to SD-related specialty care (fertility team, urology, gynecology/oncology, sexual health clinic, endocrinology, etc.).

#### Implementation

Mixed methods will be used to assess RE-AIM outcomes in a multistage design, with both explanatory sequential and convergent steps (Figure 2) [44]. Quantitative data collection will include patient and provider participant surveys assessing reach, adoption, implementation (fidelity, feasibility, acceptability, appropriateness), as well as fidelity checklist observation by the research team (S2 Appendix). EHR abstraction will be used to measure reach, adoption and representativeness (Table 4). Qualitative data will include patient and provider responses to open-ended survey questions, tracking of adaptations, observation by the research team, and nested explanatory sequential provider interviews further exploring implementation, barriers, and facilitators. Interview guides will be informed by CFIR 2.0 and will be modified as needed based on survey results; a preliminary guide is available in S2 Appendix. Adaptations to the SD screening intervention and to the implementation plan for each site will be tracked using the

**Figure 2.**
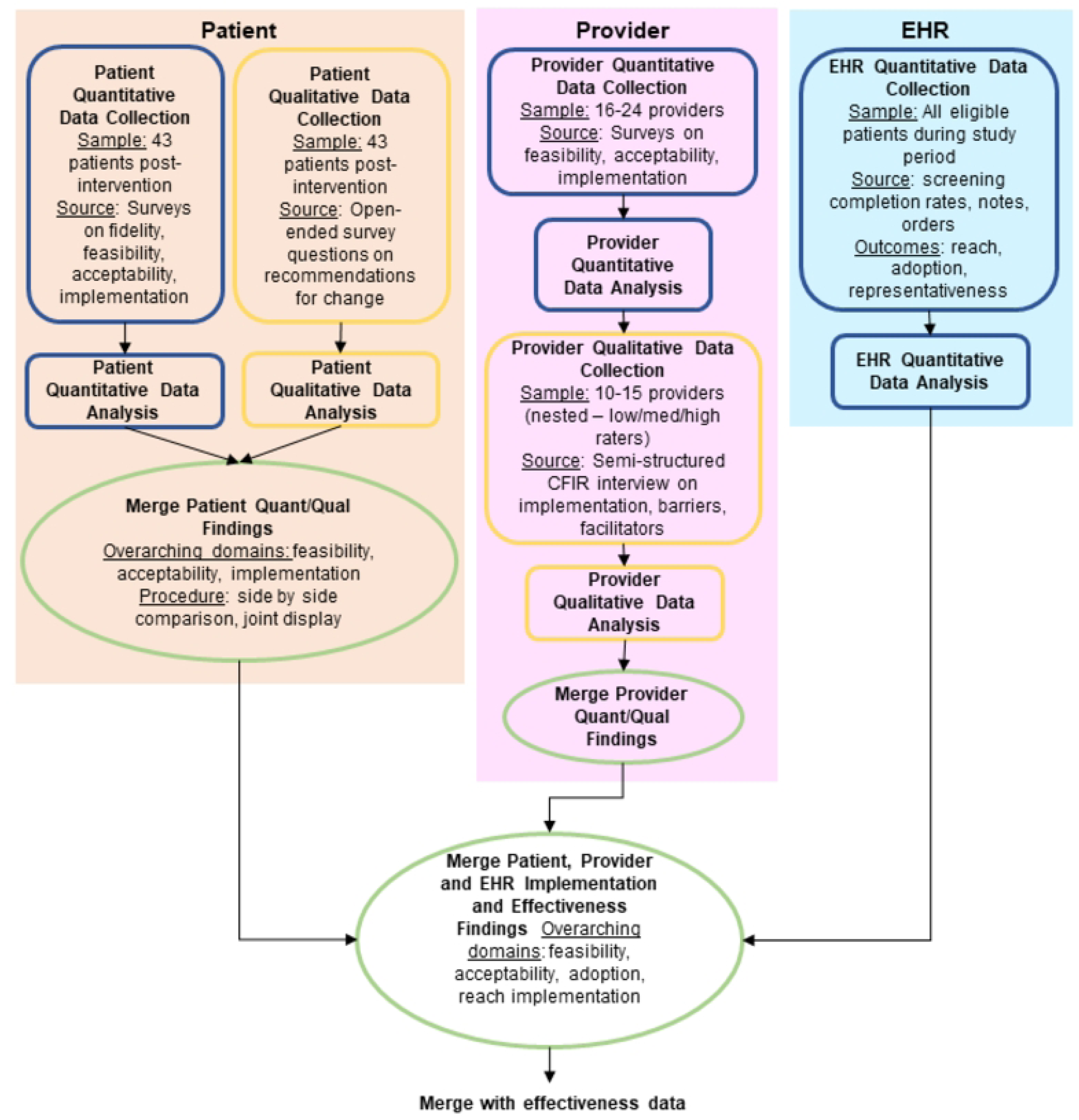
Mixed Methods evaluation of implementation of a sexual function screening intervention using patient and provider stakeholder input.

FRAME (Framework for Reporting Adaptations and Modifications-Expanded) and FRAME-IS (Framework for Reporting Adaptations and Modifications to Evidence-based Implementation Strategies), which are frameworks to characterize modifications to interventions (type of modification, reason for modification, timing, etc.) [45, 46].

### Study timeline

Pre-intervention phase study recruitment began at Children’s Hospital Colorado in February 2023 and at Seattle Children’s Hospital in December 2023, and is ongoing. The SD screening intervention phase is expected to commence in Fall 2024 after completion of ongoing iterative refinement of the prototype.

### Analysis

#### Effectiveness

Descriptive statistics (means, standard deviations, medians, ranges, frequencies, percentages) will be computed for patient characteristics, overall and stratified by study period. Differences in patient characteristics between study periods will be assessed using chi-squared and t-tests (or appropriate bivariable tests based on variable type and distribution).

The primary analysis will compare the effect of the SD screening intervention on the proportion of off-therapy AYA-CCS that self-report SD screening. We will use a multivariable logistic regression model for the binary outcome of self-reported SD screening, controlling for patient-level predictors identified a priori to be potential confounders (e.g., age, gender, presence of SD concerns, endocrinopathies, clinic site, treatment status). Although there is correlation due to the hierarchical structure of the data (patients within sites), due to the limited number of sites (<5), we will include a fixed effect for site rather than a random effect. Similar exploratory analysis will occur for on-therapy participants. We will perform a subgroup analysis by key variables (site, treatment status) using an interaction between the subgroup variable and the intervention period. The secondary outcome of whether a patient reported that their SD need was met will be analyzed similarly, overall and in the subgroup of those that indicated that they had a SD need, which will be assessed using an interaction with the SD need indicator and the pre-/post-indicator.

We will also use similar multivariable regression models adjusted for the same covariates to assess the effect of the intervention on secondary and exploratory outcomes: patient satisfaction, feasibility of collecting EHR data, and number of referral patterns. The specific link function for each regression model will be chosen appropriately based on the outcome type.

Goodness-of-fit tests and model-fitting diagnostics will be performed for proposed analyses to assess for influential points, outliers, and to evaluate alternative model specifications. All hypothesis tests will be two-sided with alpha=0.05, and p-values and confidence intervals will be reported. Statistical analyses will be conducted using R or SAS version 9.4 (SAS Institute Inc., Cary, N.C.).

#### Implementation

Because this is a pilot type 1 hybrid trial, analyses of implementation outcomes and contextual factors are descriptive, aiming to inform a future multicenter randomized hybrid trial. Mixed methods integration will occur over several steps using connecting, building, merging and embedding approaches (Figure 1) [44]. In merging steps, quantitative and qualitative survey data will be merged using a joint display [47]. Then, merged survey results will inform qualitative data collection, including nested sampling and adaptation of the CFIR-informed interview guide. Interviews will be analyzed using thematic content analysis, using both inductive and deductive coding following CFIR 2.0 [48]. We will follow a team-based, inductive process in which two team members will independently review and identify codes for the first several interview transcripts. The research team will review respective code lists and transcripts, reconciling discrepancies in code definitions and applications. Iteration will continue during data collection until the research team is calibrated and a final codebook is established. Subsequently, a minimum of 20% of transcripts will be double-coded. ATLAS.ti 24 qualitative data management software will be used. Complete qualitative and quantitative results will then be summarized in a joint display to support development of implementation strategies to strengthen future testing and delivery of the screening approach [47].

### Quality assurance

This study aims to measure preliminary effectiveness of a flexible SD screening intervention. Our sample size was selected to ensure ability to detect a clinically meaningful improvement in SD communication. By assessing the intervention across multiple clinics in two separate hospitals, the external validity of our findings will be strengthened; however, this is still considered a pilot study and assessment across more sites with varied cultural settings will be required in future studies. Further, we will use purposeful sampling to achieve maximum variation across demographic groups and sites, though future research will be required to specifically evaluate effectiveness and implementation by subgroups (e.g., gender, race, sexual orientation), and to evaluate the intervention in populations with a language preference other than English.

In this type 1 hybrid trial, use of established implementation science frameworks (RE-AIM, CFIR 2.0, FRAME, FRAME-IS) strengthens our study design. Use of mixed methods to assess implementation outcomes is expected to yield rich data to inform future implementation and dissemination. Quantitative implementation data will use validated tools for provider partners (Acceptability of Implementation Measure (AIM), Feasibility of Intervention Measure (FIM), Implementation Appropriateness Measure (IAM) increasing internal and external validity. Patient-reported acceptability, feasibility, and appropriateness will be measured with study-specific, atient-focused questions following the AIM, FiM, and IAM format [49].

All study staff have undergone training to ensure fidelity across sites. Data will be managed using REDCap and a data management plan will be used throughout the study to track data collection and ensure adherence to the study protocol [50]. Upon study completion, data will be available with request.

To increase the credibility and trustworthiness of qualitative and mixed data, we will follow the Consolidated Criteria for Reporting Qualitative Research (COREQ), with particular attention to the research team and reflexivity, study design, and analysis, and the Good Reporting of Mixed Methods (GRAMM) criteria [51, 52]. Our team-based approach, described above, will ensure analysis quality. Member checking may be implemented with a subset of participants. Use of mixed methods with multiple points of data merging, including use of quantitative data to inform qualitative interviews, also improves validity.

This protocol paper is intended to demonstrate neutrality; we will report deviations that may occur throughout the study.

### Ethics and dissemination

This trial is approved by each sites’ institutional review board (COMIRB 22-0709) and is registered at ClinicalTrials.gov (NCT05524610). This project is supported by research funds from the NIH (K08 CA263192). Informed consent will be obtained from participants prior to completing any aspect of the study. All data will be collected and stored through the password protected and secure REDCap database; audio recordings and transcripts for qualitative interviews will be stored in password protected files on the secure University of Colorado server. At each participating site, access to data will be restricted to approved study team members and study-related files will be stored in password protected files. Data will be deidentified whenever possible through the use of assigned study identification numbers. Data-sharing across sites will not include personal health information.

Findings from this study will be disseminated through presentation at academic and professional conferences and in peer-reviewed journals. Deidentified research data will be made publicly available when the study is completed and published.

## Discussion

Identification and treatment of SD in AYA CCS is an important and challenging quality of life concern. This study will determine if our patient-centered SD screening approach and implementation strategies are preliminarily effective and will describe implementation outcomes and opportunities. By developing and testing the intervention while simultaneously exploring implementation outcomes and factors, this study will inform the development of a multisite hybrid randomized trial testing clinical effectiveness and implementation strategies for the screening approach. Future studies will also employ the screening approach in a multicenter SD treatment intervention study, as well as evaluate use of the screening approach for other sensitive AYA CCS issues. Ultimately, the goal of these endeavors is to improve detection, treatment and quality of life related to SD, and other unmet concerns, in AYA CCS.

## Data Availability

Deidentified research data will be made publicly available when the study is completed and published.

## Author Contributions

**Conceptualization**: Jenna Demedis, James L. Klosky, Brooke Dorsey Holliman, Pamela N. Peterson, Christina Studts, Eric J. Chow

**Data curation:** Jenna Demedis, Julia Reedy, Kristen Miller, Junxiao Hu

**Funding acquisition:** Jenna Demedis

**Investigation:** Jenna Demedis, Julia Reedy, Kristen Miller, Junxiao Hu, Brooke Dorsey Holliman, Pamela N. Peterson, Christina Studts, Eric J. Chow

**Methodology**: Jenna Demedis, Julia Reedy, Junxiao Hu, James L. Klosky, Brooke Dorsey Holliman, Pamela N. Peterson, Christina Studts, Eric J. Chow

**Writing – Original Draft:** Jenna Demedis, Julia Reedy

**Writing – review & editing:** Jenna Demedis, Julia Reedy, Kristen Miller, Junxiao Hu, James L. Klosky, Brooke Dorsey Holliman, Pamela N. Peterson, Christina Studts, Eric J. Chow

## Supporting information captions

S1. Study Protocol: Aims 2-3 of this protocol represent the described clinical trial. Aim 1 represents a separate intervention adaptation phase.

S2. Study Appendix: Example participant surveys and interview guides

S3. Model consent forms

S4. SPIRIT Checklist

